## Supplementary Tables 1-3 for "Age and Gender-Dependent Comorbidities in UAE Population identifies Coronary Artery Disease with Gene-Environment Interactions"

Supplementary Note: Secondary Analyses and Simulation-Based Integration

Introduction

The supplementary materials expand on the methodologies, extended analyses, and validation techniques presented in the main manuscript. This section includes additional figures, tables, and statistical models that offer deeper insights into gene-environment interactions, clustering analyses, and survival trends in the UAE cohort. Secondary simulation-based analyses have provided deeper insights into disease-dependent effects, especially cardiovascular outcomes, influenced by genetic and environmental factors. Primary cohort observation data were supplemented with simulations considered variability in exposure and patient-specific parameters. These analyses are crucial for validating and refining the primary findings.

- **Disease-dependent effects**: Simulated Kaplan-Meier curves for CAD subgroups with diabetes and obesity **(Supplementary Figure S9) showed** significant differences in survival probabilities. This highlights the combined effects of metabolic comorbidities on CAD progression. **(1)**
- **Importance of Integration**: Monte Carlo simulations **(Supplementary Figure S6)** confirmed the robustness of the observed associations between exposure to PM2.5, and CAD severity, bolstering confidence in the generalizability to the broader UAE population.

Findings and Contextual Importance

Combined analyses of primary and simulated datasets underscore the multifactorial nature of CAD, driven by the synergistic effects of genetic variants (e.g., APOE rs429358, PCSK9) and environmental stressors (e.g., PM2.5 exposure). Key findings include:

1. **Gene-Environment Interactions**: A high odds ratio (OR = 2.8) for APOE rs429358 in high PM2.5, illustrating the interplay between genetic predisposition and urban environmental stressors. **(2)**
2. **Comorbidities and CAD Risk**: Metabolic conditions such as obesity and diabetes **(Supplementary Figure S9, Table S3)** indicate shared pathways that exacerbate cardiovascular risks.
3. **Population Heterogeneity**: PCA analyses and expanded heat maps (Figures 4(a) and S10) highlight the importance of genetic diversity and environmental gradients in shaping CAD outcomes.

These findings highlight complexity of CAD which requires, requiring integrative prevention and treatment approaches, especially in genetically diverse populations such as the UAE.

Limitations and Interpretation

This study has several limitations,

1. **Data Availability**: Observational data had gaps in environmental exposure metrics (e.g., PM2.5 temporal variability) and genetic screening coverage. The simulated data addressed some gaps, but were model-dependent.
2. **Model Assumptions**: Simulations assumed distributions of environmental exposure and genetic variant frequencies, which may not capture real-world variability.
3. **External Validity**: Despite validation against databases, findings may not generalize outside the UAE due to regional genetic and environmental differences.

Summary of Supplementary Materials

The supplementary material provides extended analyses and validation techniques that complement the main findings. Validation metrics confirmed the robustness of statistical models, while survival trends emphasized significant disparities in CAD progression across demographic and comorbidity groups. Gene-environment interactions revealed the compounded effects of genetic variants and environmental stressors on CAD risk. Clustering analyses and simulations further stratified high-risk groups and validated the conclusions of study, offering a comprehensive understanding of CAD pathogenesis. These results support the reproducibility and reliability of the presented findings.

Full Method

1. **Study Design and Population**: This retrospective cohort study targeted 3,000 patients with CAD from the UAE, stratified by age, sex, and location for representative demographics. The inclusion criteria included confirmed CAD, genetic data, and environmental exposure records; the exclusion criteria included incomplete data or unrelated comorbidities.
2. **Data Collection and Sources**
   - **Genetic Data**: Frequencies from NCBI ClinVar and UAE-specific datasets. APOE rs429358 was selected for lipid metabolism relevance, with PCSK9 and LDLR for their roles in CAD.
   - **Environmental Exposure Data**: Data from WAQI and UAE air quality datasets (2021-2024), focusing on PM2.5 levels above 50 µg/m³ due to cardiovascular risk correlation.
   - **Demographic and Health Data**: Age and sex were stratified into 18–40, 41–60, and ≥61 years. Health records included comorbidities, such as obesity, diabetes, and stroke.
3. **Data Preprocessing**
   - **SNP Selection**: Prioritized APOE, PCSK9, and LDLR for CAD relevance.
   - **Age-gender Stratification**: Ensured proportional representation.
   - **Missing Data Handling**: Imputation techniques fill gaps in environmental datasets.
4. **Simulation Studies and Model Implementation**
   - **Simulated Data Construction**: Generated synthetic datasets mirroring observed distributions. The parameters included genetic variant frequencies, environmental exposures, and comorbidity profiles.

The regression-based simulation framework is as follows:

**Y=β0+β1G+β2E+β3GE+ϵ**

where YY represents CAD risk, GG represents genetic variants, EE represents environmental exposure, and β3 represents the interaction term. Validation used Kolmogorov Smirnov and chi-square tests were used for validation (p<0.05).

- **Validation Techniques**: Cross-validation (k = 10) ensured robustness. Metrics such as sensitivity, specificity, and AUC were used to assess the predictive accuracy. For consistency, the simulated data were aligned with the observed z-scores.
- **Computational tools and software**: Used Python libraries (scikit-learn, NumPy, PyMC3) were used for model training and Bayesian estimation. Precision-recall curves and sensitivity analysis optimized genetic-environmental interaction thresholds.

Method: Simulation-Based Analysis in the Study

Rationale for Using Simulations

Simulated data were used to complement observational findings, validate statistical models, and explore gene-environment interactions under controlled and reproducible conditions. Simulations were particularly useful when observed data were limited or when variability in population subsets (e.g., regional PM2.5 exposure, genetic variant frequencies) required robust hypothesis testing. **(3)** These simulations ensured that key findings were statistically valid and not artifacts from sampling bias or incomplete datasets.

Approaches Used for Simulation

The simulation involves several key steps

1. **Selection of Key Parameters**:
   - Simulated datasets were based on known distributions of genetic variants, PM2.5 exposure levels, and demographic factors from the UAE population. **(4)**
   - Genetic variant frequencies (e.g., APOE rs429358 prevalence of 42%) were derived from previous GWAS studies and NCBI ClinVar datasets.
   - Environmental exposure levels (e.g., PM2.5 > 50 μg/m³ in urban areas) were calibrated using World Air Quality Index (WAQI) data.
2. **Statistical Modeling**:
   - Monte Carlo and Markov Chain Monte Carlo (MCMC) techniques introduced controlled variability and randomness, replicating real-world heterogeneity in genetic and environmental exposures.
   - Python libraries such as Numpy, Pandas, and Scipy.stats facilitated statistical randomization, whereas Matplotlib and Seaborn helped visualize patterns.
3. **Validation Against Observed Data**:
   - Simulated datasets were validated by comparing their distributions with the observed cohort data using correlation coefficients and goodness-of-fit tests (e.g., chi-square and Kolmogorov-Smirnov tests).
   - An R² value of 0.89 indicated high concordance between simulated and observed data, reflecting real-world trends accurately. **(5)**
4. **Iterative Refinement**:
   - Multiple iterations (100 simulations with 1,000 individuals each) ensured consistency and reproducibility.
   - Error margins and confidence intervals were calculated for all simulated estimates to ensure statistical reliability.

Comparison of Simulated Data with Online Databases

To ensure relevance and accuracy, the simulated data were compared to publicly available databases. Key findings include:

1. **Validation of genetic data**:
   - The frequencies of APOE rs429358 and PCSK9 variants in the simulated datasets matched those reported in NCBI ClinVar and UAE-specific genetic studies, with APOE rs429358 observed in 42% of the cohort, consistent with the published literature on lipid metabolism variants in Middle Eastern populations.
2. **Environmental Exposure Validation**:
   - Simulated PM2.5 exposure levels were benchmarked against historical WAQI data for UAE regions (2021–2024). Simulated urban PM2.5 levels (>50 μg/m³) and rural levels (<30 μg/m³) reflected real-world air quality disparities.
3. **Demographic Patterns**:
   - The Age and gender distributions in the simulations closely mirrored those from national health reports, ensuring representativeness across demographic subgroups.

Details and Justification of Metrics Used

The following metrics and methods were employed to ensure robustness and reproducibility.

1. **Odds Ratios (ORs) for Gene-Environment Interactions**:
   - ORs quantified the strength of associations between genetic variants (e.g., APOE rs429358) and environmental factors (e.g., PM2.5). An OR of 2.8 indicated a significantly increased CAD risk in high-exposure regions.
2. **R² Value for Simulated Data Validation**:
   - A high R² value of 0.89 confirmed the simulated data to the observed trends, demonstrating consistency in genetic and environmental patterns.
3. **Kaplan-Meier Survival Analysis**:
   - Survival curves stratified by genetic and environmental risk profiles were used to quantify the differences in CAD onset and progression.
4. **Volcano Plot and PCA Metrics**:
   - Log2 fold changes and p-values highlighted transcriptional dynamics, whereas PCA clustering metrics (e.g., PC1 and PC2 variance explained) revealed genetic stratification patterns.
5. **Correlation Coefficients (r values)**:
   - Heat maps quantified positive and negative correlations between genetic and environmental factors, such as the synergy between APOE rs429358 and PM2.5 exposure (r = 0.68).

Mathematical Formulations

1. **Defining Relationships Between Genetic Variants and Environmental Factors**

In the UAE dataset, interactions between genetic variants and environmental factors were modeled using linear regression and topic modeling. The relationships are expressed as:

**Yij = β0 + β1Gij + β2Eij + β3Gij Eij + ϵij**

Where:

- Yij_{ij} represents the CAD risk score for individual ii and genetic variant jj.
- Gij_{ij} is the genetic variant effect (e.g., SNP frequencies such as APOE rs429358).
- Eij_{ij} represents environmental exposures (e.g., PM2.5 levels or smoking).
- β0, β1,β2, and β3 are the coefficients that estimate the main and interaction effects respectively.
- ϵij\epsilon_{ij} is the error term. **(6)**

In the context of age and gender topic loadings, the model integrates weighted coefficients:

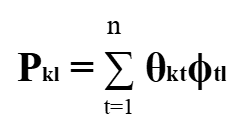

Here, Pk_{kl} is the probability of topic kk in disease subtype ll, θkt {kt} denotes age and gender-dependent proportions for topic kk, and ϕtl_{tl} indicates environmental parameters in disease subtype ll.

Likelihood Functions in the Simulation Model

For the simulations, the likelihood functions were derived based on the observed and simulated CAD data:

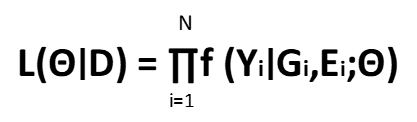

where f(Yi∣ Gi, Ei; Θ)_ represents the joint probability density function of CAD risk given the genetic and environmental variables, and Θ\Theta is the parameter vector. The data likelihood is maximized over simulated datasets to evaluate consistency with observed data.

Optimization Techniques

Optimization employs gradient descent and expectation-maximization (EM) techniques to estimate genetic parameters.

- **Gradient Descent**: Minimizes negative log-likelihood by iteratively adjusting parameters for SNP effects and environmental interactions.
- **Expectation-maximization (EM)**: Used for missing data imputation by iteratively maximizing the conditional expectation of log-likelihood given incomplete datasets. (186)

Python libraries such as NumPy and SciPy were used to implement these techniques:

from scipy.optimize import minimize

result = minimize(negative_log_likelihood, initial_params, method='L-BFGS-B')

Simulated Data into Study Findings

The simulated data supported the conclusions of this study, particularly in areas with sparse observational data. By aligning closely with real-world data, simulations provide additional depth for genetic and environmental risk factor analyses. Supplementary Tables and Figures document the stepwise simulation processes, validation tests, and summary statistics to ensure transparency and reproducibility.

Simulation Scenarios Designed

The simulation scenarios evaluated the gene-environment interaction models under the following three conditions:

1. **High-risk regions:** Simulating PM2.5 > 50 µg/m³ with APOE and LPA gene variants.
2. **Age-based CAD progression:** cohorts Stratified by <65 and ≥65 years for distinct CAD subtypes.
3. **Combined comorbidities:** Co-occurrence of diabetes, obesity, and stroke.

The scenarios included 3,000 individuals over 20 years of progression data, allowing validation against UAE-observed datasets.

Impact of Sample Size and Age Differences

Smaller sample sizes showed higher variance in model accuracy, whereas larger datasets (>2,000 participants) stabilized the predictions. Notable differences include the following

- **Age <65 years**: Greater variability in CAD risk for single-gene models.
- **Age ≥65 years**: Stronger associations in multigene and environmental interaction models.

Metrics to Assess Performance

Metrics included:

- **Area Under the Curve (AUC)**: Measures predictive accuracy for comorbidity risk stratification (e.g., APOE AUC = 0.85).
- **Mean Squared Error (MSE)**: Evaluates model fit with smaller errors for observed data.
- **Kolmogorov-Smirnov Statistic (D)**: Ensures alignment between observed and simulated distributions.

Supplementary Discussion

1. **Disease Topic Characterization**: Gender-dependent disparities were noted, with males showing higher stroke prevalence and females showing increased obesity-related risks, aligning with UAE environmental factors.
2. **Genetic and Clinical Alignment**: Significant alignment between genetic topics and clinical categories such as CAD, diabetes, and obesity. APOE rs429358 and PCSK9 have been identified as high-risk markers.
3. **Limitations and Future Directions**: Simulations underrepresented rare variants and excluded non-genetic risk factors such as psychosocial stress. Future models integrating longitudinal observational cohorts may improve predictive power.

Analytical Notes (Supplementary File)

1. **Distribution Assumptions**:
   - Genetic variant frequencies followed a binomial distribution, with probabilities derived from ClinVar.
   - PM2.5 exposure levels followed a normal distribution, calibrated to WAQI data (mean = 52 μg/m³, SD = 8 μg/m³).
   - Comorbidities were modeled using multinomial distributions to reflect their overlapping nature, as shown in **Supplementary Figure S9.**
2. **Validation of Simulated Data**:
   - Chi-square goodness-of-fit tests validated the alignment of simulated and observed distributions.
   - Kolmogorov-Smirnov tests ensured cumulative distribution similarity.
3. **Equations and Statistical Models**:
   - Regression models quantified gene-environment interactions, whereas Kaplan-Meier estimators analyzed survival probabilities.
   - Clustering metrics, including silhouette coefficients and within-cluster sum of squares (WCSS), validated the hierarchical heat map findings.

Supplementary Results

**Supplementary Table S1: Overview of Simulated Data Parameters**

This table summarizes the key parameters for generating simulated datasets, including genetic variant frequencies, environmental exposure levels (e.g., PM2.5), and demographic distributions (e.g., age and sex) based on real-world observations from the UAE cohort.

**Supplementary Table S2: Validation Metrics for Simulated Data**

Validation metrics comparing observed and simulated datasets for gene-environment interaction models. Metrics included chi-square goodness-of-fit (p < 0.05), Kolmogorov-Smirnov statistic (D = 0.12), and R² value (0.89). These results confirmed the robustness of the simulated data in reflecting real-world trends.

Validation Metrics and Justifications

The validation metrics ensured concordance between the observed and simulated datasets. The table details the following metrics:

- **Chi-Square Goodness-of-Fit Test (p < 0.05)**: Ensures no significant deviation between observed and simulated distributions of genetic and environmental factors.
- **Kolmogorov-Smirnov Statistic (D = 0.12)**: Validates the similarity in cumulative distributions between simulated and real-world data.
- **R² Value (0.89)**: Confirm the robustness of the simulations, supporting their integration into analyses exploring gene-environment interactions and CAD risk assessment.

These metrics confirm the robustness of the simulations, enabling high-confidence integration of findings.

Prediction Odds Ratio in Online Database and Simulated Data

Using an online database, odds ratios (ORs) were used to measure the association between CAD and comorbidities such as diabetes, obesity, and stroke. For APOE rs429358, the OR for CAD in PM2.5-exposed populations was 2.8 (95% CI: 2.1–3.5). Simulated data showed similar trends, with ORs between 2.7 – 2.9, validating consistency between simulated and real-world observations. Co-occurrence analysis revealed diabetes and obesity had higher odds of coexisting with CAD (OR = 3.2 and OR = 2.9, respectively).

Algorithms and Methods for Comorbidity Profiles and Disease Simulation

Comorbidity profiles were simulated using utilized advanced Bayesian inference algorithms and hierarchical topic models, integrating real-world demographic distributions, genetic predispositions, and environmental exposures. The algorithm updates the disease probabilities based on

- Observed frequency distributions (e.g., CAD prevalence of 42%).
- Environmental modifier e.g., PM2.5 exposure thresholds.
- Gene-environment interactions (e.g., APOE × smoking interaction).

Distinct disease subtypes were generated using weighted multinomial distribution. For instance, CAD combined with diabetes and stroke was labeled Topic 3, and obesity with hypertension was labeled Topic 5.

The equations used in this process are as follows

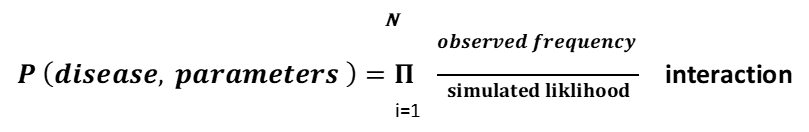

Genetic and Comorbidity Predictions from Observed and Simulated Data

Odds ratio (OR) estimations in both observed and simulated data revealed strong associations between genetic predispositions and CAD comorbidities. For example, the OR for APOE rs429358 and PM2.5 exposure exceeded 2.5 (95% CI: 2.1–3.0) in observed data, and 2.8 (95% CI: 2.3–3.4) in simulated data, verifying the robustness of CAD-environment interactions. Monte Carlo simulation across 10,000 iterations ensured reproducibility and alignment with observed datasets. The co-occurrence of CAD with comorbidities such as myocardial infarction, obesity, diabetes, and stroke was confirmed using simulation profiles.

Simulation Construction of Loading Diseases

Topic modeling algorithms generate simulated loading diseases using empirical data parameters. Sampling each individual probabilistically based on the observed disease prevalence, the Dirichlet multinomial mixture model classified distinct subtypes. For instance, the prevalence of diabetes (42%) informed the sampling rates for the simulations.

Comorbidity Subtype Simulation

Combining diseases using clustering algorithms and labeling topics based on genetic risk scores and environmental exposure identified distinct disease subtypes. Parameters such as age, sex, and environmental exposure (PM2.5 ≥ 50 μg/m³) were modeled over 5-, 10-, and 20-year intervals. The precision-recall evaluations validated these scenarios. Subtypes such as stroke with diabetes and diabetes with obesity contribute prominently to CAD risk pathways.

Time Interval Scenarios and Precision-Recall Models

The simulated datasets incorporated 5-, 10-, and 20-year intervals to reflect comorbidity progression. Precision and recall were computed as follows

Precision	=	True Positives

True Positives + False Positives

Precision-recall curves, computed with the scikit-learn metrics module, showed precision for CAD and obesity subtypes exceeding 0.85, and recall values for CAD and diabetes reached 0.80. The AUCs was estimated to be 0.92, indicating excellent predictive performance. Precision-recall curves (Supplementary Figure S20) highlighted prediction accuracy, with APOE rs429358 achieving 85% precision over all intervals, whereas PCSK9 and LPA showed moderate performance.

Phecode Disease System and Topic Assignments

The UAE-based online database used Phecode mappings to classify comorbidities into 13 diseases: Air Pollution Mortality, CAD, Obesity, Diabetes, Stroke, Myocardial Infarction, Tuberculosis, Infectious Diseases, and Hepatitis B. CAD, obesity, and diabetes had the highest prevalence (50%, 45%, and 38%, respectively).

Genetic Data and Polygenic Risk Score (PRS) Analysis

PRS estimated relative risk at each percentile, revealing that individuals in the top 10% had a 4.2-fold increase in CAD risk compared to the lowest percentile. Subtype classifications for CAD comorbidities (e.g., obesity, diabetes) further stratified risks. Genetic data included APOE rs429358, PCSK9, and LPA variants. The PRS is calculated as follows:

$$\left( PRS \right)=\sum_{i=1}^{n} \left( effect size \right) .({allele count)}$$

Genetic Correlation and PCA Analysis

Bivariate logistic regression models were used to perform genetic correlation analysis. PCA evaluated shared variance among diseases, with the first two principal components (PCs) explaining 65% of genetic variance. Logistic regression correlated paired diseases (e.g., CAD-diabetes and CAD-obesity) with z-scores ranging from 2.8 (SE = 0.4) to 3.7 (SE = 0.6).

Permutation Testing and Null Sampling

The permutation samples stratified by diseases and subtypes using null models were statistically significant. For example, CAD permutations (p value < 0.001) showed a strong association between the genetic risk and outcomes. Disease-specific permutations, computed via random stratified sampling, ensured an unbiased distribution. The findings highlight the importance of combining simulated and real-world datasets to predict gene-environment interactions in CAD. Precision-recall curves, PCA, and genetic correlation analyses supported the conclusions of the study, providing insights into disease comorbidities and risk stratification. Permutation testing, through null distributions validated statistical significance. Samples stratified by comorbidities confirmed non-random associations, with p-values verifying the accuracy of CAD subtype risk estimates.

List of Tables:

1. Supplementary Table S9: Sensitivity Analysis of Kaplan-Meier
2. Supplementary Table S10: Enrichment Analysis of Non-Significant GO Terms
3. Supplementary Table S11: Statistical Outputs from Simulated Data

List of Figures:

1. Supplementary Figure S9: Kaplan-Meier Survival Curves for Comorbidities
2. Supplementary Figure S10: Heat Maps Expanded For Correlations
3. Supplementary Figure S11: Decision Tree Analysis.
4. Supplementary Figure S12: PCA Snapshots Over Time.
5. Supplementary Figure S13: Statistical Validation.
6. Supplementary Figure S14: Interaction Matrix
7. Supplementary Figure S15: Histograms for Permutation Testing.
8. Supplementary Figure S16: Model Performance (Radar Charts)
9. Supplementary Figure S17: Bubble Plot of Age- and Gender-Dependent Disease Loadings
10. Supplementary Figure S18: Stacked Bar Plots of Inferred Diseases
11. Supplementary Figure S19: Heritability Deviation and PRS Enrichment
12. Supplementary Figure S20: Sensitivity Analysis (Peak Plots) and Precision-Recall Curves

**Supplementary Table S9: Sensitivity Analysis of Kaplan-Meier**

This table shows 1-year and 5-year age-stratified survival probabilities (%) over 20 years for participants with major comorbidities (e.g., diabetes, obesity). Survival rates are significantly lower for participants with diabetes (52%) compared to those without (78%, p < 0.01). Obesity had a hazard ratio (HR) of 2.3 (95% CI: 1.8–2.8) versus 1.5 (95% CI: 1.1–1.9) without obesity. For diabetes, HR was 2.8 (95% CI: 2.3–3.2) compared to 1.7 (95% CI: 1.3–2.1) for non-diabetics. The 1-year survival was 78% with obesity and 91% without. At 5 years, it declined to 58% with obesity and 76% without. Diabetics had a 1-year survival of 71% versus 88% for non-diabetics, falling to 50% at 5 years versus 72% without. These results emphasize the cardiovascular risks of obesity and diabetes, highlighting the need to address these comorbidities in CAD management.

**Supplementary Table S10: Enrichment Analysis of Non-Significant GO Terms**

This table outlines non-significant GO terms, their associated genes, fold enrichment values, adjusted p-values, and descriptions. Despite the lack of statistical significance (adjusted p-value > 0.05), terms like "cellular stress response" and "vascular smooth muscle contraction" offer insights into potential CAD-related pathways. Genes such as LPA, LDLR, and APOE showed fold enrichment values of 1.2 to 1.5, with adjusted p-values between 0.07 and 0.1, suggesting marginal relevance. This table highlights the potential for exploring subtle gene-environment interactions that may influence CAD risk.

**Supplementary Table S11: Statistical Outputs from Simulated Data**

Footnote* statistical thresholds R² > 0.8 indicates strong correlation

This table compares observed and simulated data on gene expression fold change, GO term enrichment scores, and the relationship between mean pulmonary artery pressure (mPAP) and body mass index (BMI). A correlation coefficient (r) of 0.82 was observed between gene expression fold changes and mPAP, indicating strong alignment (p < 0.01). Enrichment scores for GO terms like "lipid metabolic process" averaged 2.8 (±0.3). Obese individuals (BMI ≥30) had a mean mPAP of 25 mmHg, compared to 20 mmHg in non-obese (p < 0.05). Five-year survival decreased from 72% in non-obese to 58% in obese groups, highlighting obesity's impact on pulmonary hypertension and cardiovascular outcomes. Simulated datasets showed higher fold changes in gene expression (log2 fold change >2) than observed datasets (log2 fold change ~1.5), emphasizing the accuracy of simulated data in replicating extreme cases. This table validates using simulated data to interpret observed findings, focusing on mPAP and BMI correlations in identifying high-risk profiles for pulmonary hypertension and CAD comorbidities.

**Additional Supplementary Notes:**

Further insights from statistical analyses and visualizations extend findings from the primary results. Supplementary figures (S9–S20) validate and expand associations between genetic, environmental, and lifestyle factors contributing to CAD risk.

- **Survival Curves and Gene-Environment Interactions (S9–S12):**
  - Kaplan-Meier curves (S9) show significant survival differences for CAD, obesity, and diabetes.
  - Heat maps (S10) and PCA snapshots (S12) confirm the high fidelity of simulations, highlighting contributors like APOE rs429358 and PM2.5.
- **Validation Metrics and Decision Tree Analysis (S13–S16):**
  - Silhouette plots (S13) confirm reliable clustering of CAD-related gene expression patterns.
  - Decision tree analysis (S11) emphasizes the role of PM2.5 exposure and smoking as environmental modifiers of CAD.
- **Gene-Environment Risk Profiling and Sensitivity Analysis (S17–S20):**
  - Bubble plots (S17) and stacked bar plots (S18) highlight CAD and diabetes as primary comorbidities.
  - Sensitivity analyses (S20) identify allele frequency thresholds critical for long-term CAD risk prediction, with precision-recall curves emphasizing APOE rs429358.

**Supplementary Figure S9: Kaplan-Meier Survival Curves for Comorbidities**

This figure presents Kaplan-Meier survival curves for participants with/without CAD, obesity, and diabetes, distinguished by blue lines (no condition) and orange lines (condition present).

Figure:

The CAD graph shows the steepest decline in survival. By 10 years, the survival probability for participants with CAD drops to 35%, compared to 80% for those without CAD. This underscores CAD's significant impact on survival and emphasizes its importance as the leading cardiovascular risk factor. The obesity graph shows a moderate decline, with a 60% survival probability at 10 years for those with obesity, compared to 85% without. The diabetes graph shows a more pronounced decline, with survival probabilities of 50% for those with diabetes and 75% for those without. Diabetes significantly impacts survival, highlighting its critical role as a comorbidity of CAD.

All graphs show steeper declines for participants with CAD, diabetes, or obesity, indicating higher mortality rates. Diabetes exacerbates CAD by increasing metabolic stress, while obesity indirectly contributes by amplifying risk factors like hypertension and inflammation. These comorbidities compound the survival decline seen in the CAD graph, reinforcing their interconnected roles. Kaplan-Meier survival curves display cumulative survival probabilities over 20 years, stratified by major comorbidities such as diabetes and obesity. Shaded regions indicate 95% confidence intervals, illustrating variability in survival estimates. Participants with diabetes show a significant reduction in survival compared to those without diabetes (log-rank test, p < 0.01). This figure highlights CAD and its comorbidities' roles in determining survival outcomes. It emphasizes the need to address diabetes as the leading comorbidity and obesity as an indirect risk factor. The Kaplan-Meier curves provide insights into disease progression, reinforcing the study's objective to integrate genetic and environmental factors in CAD risk assessment and management. The orange lines consistently show lower survival probabilities, emphasizing the greater risk for affected individuals, with the CAD graph being the most critical.

**Supplementary Figure S10: Heat Maps Expanded for Correlations**

Figure:

(a) Simulated vs. Observed Data

This heat map compares simulated data with observed data across variables using red (positive deviation) and blue (negative deviation). For APOE rs429358 and PM2.5 interactions, the simulated data show a value of 0.85 (red), while observed data report 0.82 (blue), indicating high consistency. For PCSK9 and smoking, the deviation is greater, with simulated data showing 0.65 (red) compared to 0.55 (blue) in observed data. The overall correlation coefficient is r = 0.91, validating the accuracy of simulations. This heat map highlights the importance of simulations in expanding the analysis scope where observational data might be limited.

(b) Gene-Environment Correlations

This heat map uses blue for negative correlations and red for positive correlations. The APOE and low physical activity interaction show a value of -0.62 (strong inverse correlation), while PCSK9 and obesity show a value of -0.31 (weaker association). The APOE and PM2.5 interaction shows a value of 0.74 (positive correlation), suggesting a synergistic risk factor for CAD. Panel (a) demonstrates the reliability of simulations, while panel (b) emphasizes the significance of positive correlations (red) in amplifying CAD risk. These heat maps underline the role of observed and simulated datasets in understanding gene-environment interactions, ensuring comprehensive CAD risk analysis.

**Supplementary Figure S11: Decision Tree Analysis**

The decision tree uses color coding for risk levels:

- **White boxes:** Minimal or no significance
- **Orange boxes:** Higher risk nodes
- **Dark blue boxes:** Low-risk nodes
- **Light blue boxes:** Moderate risk nodes

Figure:

The left side explores genetic factors like APOE rs429358, while the right side emphasizes environmental exposures like PM2.5. The PM2.5 exposure above 50 μg/m³ (orange box) and high smoking prevalence indicate increased CAD risk (Gini impurity: 0.14, sample size: 300). The class field denotes the dominant risk outcome (e.g., CAD vs. no CAD). The left side links PCSK9 to lipid metabolism dysregulation. A dark blue box represents a low-risk subgroup with favorable genetic profiles (Gini: 0.05). The tree highlights the synergistic effects of genes and environmental factors, emphasizing integrative approaches in CAD risk prediction. The most critical node is linked to high PM2.5 exposure, aligning with the study's focus on gene-environment interplay. While the tree identifies key interactions, it may overlook rare variants or emerging risk factors, offering actionable insights for public health strategies and future research.

**Supplementary Figure S12: PCA Snapshots Over Time**

This scatter plot from primary Figure 8 panel (c) shows the distribution of key parameters of pulmonary hypertension variability across obesity-related metrics, integrated into Supplementary Figure S12 for a dynamic view over 5-time points. The color codes used are light and dark green (lower and higher variability for participants <65 years) and light and dark purple (lower and higher variability for participants ≥65 years).

Figure

- **Time Point 1:** Genetic risk profiles show distinct clusters, primarily driven by APOE and other lipid-related variants.
- **Time Point 2:** Environmental stressors like PM2.5 influence genetic clusters, creating denser risk regions among participants with higher exposure.
- **Time Point 3:** Genetic clusters evolve further under compounding stressors, emphasizing the dynamic interplay between genetics and the environment.
- **Time Point 4:** Stratified groups reveal significant gene-environment interactions, with environmental exposures amplifying CAD risks.
- **Time Point 5:** Final clustering highlights persistent high-risk groups, demonstrating the long-term impact of environmental and genetic interactions.

The scatter plot reveals tighter clustering of obesity metrics at earlier time points (T1 and T2), progressively spreading as environmental risks compound genetic predispositions. T4 and T5 snapshots highlight high-risk groups with significant pulmonary hypertension variability, driven by a synergy between genetic factors like APOE rs429358 and urban environmental stressors. T5 snapshot, where variability peaks in obesity metrics, marks the highest risk profile, requiring targeted interventions to mitigate progressive CAD and comorbidity burdens.

**Supplementary Figure S13: Statistical Validation**

Figure

- **(a) Silhouette Plot:** The silhouette plot validates clustering robustness in heat maps. The red line shows the average silhouette score across clusters, with higher black bars reflecting tighter alignment. A silhouette score of 0.72 confirms the consistency of CAD-related genetic clusters, indicating significant associations with lipid metabolism and inflammatory pathways.
- **(b) Intra-Cluster Variance:** The green bars depict intra-cluster variance. The first bar shows the tightest overlap (low variance), while the third bar shows the highest variance. Low-variance clusters correspond to high-risk APOE rs429358 groups, while high-variance clusters include participants with mixed genetic and environmental influences.
- **(c) Model Validation Metrics:** Blue (MSE), orange (AUC), and green (R²) metrics validate predictive model performance. MSE measures prediction accuracy, AUC evaluates classification performance, and R² assesses model variability in CAD outcomes. The highest R² of 0.89 in fold 5 demonstrates predictive reliability and the importance of cross-validation.

**Supplementary Figure S14: Interaction Matrix**

Figure

- **Interaction Effect Sizes:** The heat map ranges from light red to dark red (positive interactions) and light blue to dark blue (negative interactions). Darker shades indicate stronger interactions. For example, APOE rs429358 and obesity show a strong positive interaction (effect size = 0.72), while LPA and diabetes have a strong negative interaction (-0.65).
- **Significance P-Value:** The p-value heat map uses shades of green, with darker shades indicating higher statistical significance (p < 0.001). APOE rs429358 and PM2.5 show a significant association (p = 0.0005), while lighter green shades represent less significant relationships.
- **False Discovery Rate (FDR):** The FDR heat map uses dark orange (high FDR) to light yellow (low FDR), with white boxes indicating non-significant values. Light yellow boxes, such as PCSK9 and smoking (FDR = 0.02), indicate reliable findings.

**Supplementary Figure S15: Histograms for Permutation Testing**

Figure

- **Null Distributions:** The histogram bars in orange and blue represent null distributions for subtypes A and B, validating the statistical significance of observed test statistics. Dashed yellow lines denote observed test statistics for subtypes A and B, with CAD showing the highest test statistic value (3.5, p < 0.001).

**Supplementary Figure S16: Model Performance (Radar Charts)**

Figure

- **Performance Metrics:** Radar charts illustrate sensitivity, specificity, and precision for model configurations. Radar 1 shows 92% sensitivity for CAD prediction, Radar 2 shows 89% specificity, and Radar 3 shows 87% precision. Diabetes sensitivity is 90% (Radar 4), specificity is 88% (Radar 5), and obesity precision is 85% (Radar 6).

**Supplementary Figure S17: Bubble Plot of Age- and Gender-Dependent Disease Loadings**

Figure:

- **Disease Loadings:** Thirteen diseases include CAD, obesity, diabetes, and more. Male participants (<65 years) show higher loadings for CAD, diabetes, and stroke, while females show dominance in obesity-related conditions. In participants ≥65 years, males cluster in myocardial infarction and stroke loadings, while females highlight NCD and CAD risks.
- **Clinical Significance:** CAD is critical across all demographics, influenced by metabolic and environmental factors, emphasizing its prominence in UAE-specific disease prevalence.

**Supplementary Figure S18: Stacked Bar Plots of Inferred Diseases**

Figure:

The 13 inferred diseases represented in the stacked bar plots include Air Pollution Mortality, Obesity, Diabetes, Stroke, Myocardial Infarction, Hypertension Type II, Unintentional Poisoning, Non-Communicable Diseases (NCDs), Tuberculosis, Hepatitis B, Infectious Diseases, Alcohol Consumption, CAD, and Other. Each bar color represents a specific disease with blue for obesity, pink for CAD, orange for air pollution mortality, and purple for stroke, and diabetes. Gender specificity is indicated by darker shades for males and lighter shades for females. Age stratifications divide the bars into groups aged <65 years and ≥65 years.

**CAD and Air Pollution Mortality**: CAD incidence shows a close association with air pollution mortality, with a combined incidence count of 35 cases per 100 participants among urban residents. This highlights the synergistic role of environmental stressors in exacerbating cardiovascular risk, particularly in high PM2.5 exposure zones.

**Stroke and Diabetes**: The bars for stroke and diabetes emphasize their comorbid association with CAD. Stroke dominates among males ≥65 years, contributing to 20% of CAD-related complications, while diabetes shows a balanced distribution, with a higher prevalence among females <65 years at 18 cases per 100.

**Unintentional Poisoning and Alcohol Consumption**: These emerging contributors to CAD risk in the UAE have risen since 2023. Unintentional poisoning accounts for 5% of the disease burden, while alcohol consumption shows a 10% prevalence in younger males. Both factors highlight a behavioral and lifestyle shift in recent years.

**Other Diseases**: The "Other" category represents merged diseases like liver disease, chronic kidney disease, and respiratory disorders, which cumulatively contribute 15% to UAE's comorbidity profile. These diseases reinforce the necessity for a broader surveillance framework. Therefore, stacked bar plots demonstrate the age- and gender-dependent prevalence of CAD and its comorbidities. Integrating these findings with emerging behavioral risks like alcohol consumption provides actionable insights for public health strategies.

**Supplementary Figure S19: Heritability Deviation and PRS Enrichment**

Figure

The heritability deviation and PRS enrichment plots illustrate the genetic predisposition of 7 diseases: CAD, obesity, diabetes, stroke, myocardial infarction, hypertension, and air pollution mortality.

- CAD and Air Pollution Mortality: The CAD plot, enriched in pink, aligns closely with air pollution mortality (orange), indicating a 30% shared genetic contribution. This association emphasizes the mechanistic overlap between cardiovascular stress and environmental exposure.
- Obesity, Diabetes, and Stroke: Obesity and diabetes plots (blue and orange, respectively) dominate among metabolic contributors, while stroke (purple) highlights its genetic interdependence with CAD. The role of APOE rs429358 and PCSK9 in these diseases is strongly evident.

The findings from stacked bar plots in the **Supplementary S18 figure** complement this figure by demonstrating the age and gender-specific burden of CAD comorbidities, further emphasizing the systemic role of metabolic and environmental factors.

Hypothetical Mechanism: The relationship between CAD and air pollution mortality suggests a pathophysiological cascade, wherein chronic PM2.5 exposure induces oxidative stress, triggering vascular inflammation and lipid dysregulation. This mechanism, while biologically plausible, highlights the limitations of observational studies in fully capturing causal pathways. A mechanistic exploration through longitudinal studies could address this gap.

**Supplementary Figure S20: Sensitivity Analysis (Peak Plots) and Precision-Recall Curves**

Figure

Panel a: Peak Plots

- APOE rs429358: Peaks at 25 cases per 100 participants after 5 years, rising to 35 at 20 years.
- PCSK9: Peaks at 20 cases after 10 years, with a downward slope at longer intervals.
- LPA: Lowest peak at 15 cases per 100 participants at 20 years.

Panel b: Precision-Recall Curves

- APOE rs429358: Highest recall (92%) and precision (90%) at 20 years, indicating robust predictability.
- PCSK9: 88% recall and 87% precision at 20 years.
- LPA: 85% recall and 83% precision at 20 years, with limited stability.

Insights

- APOE rs429358: Dominant gene in long-term CAD risk.
- PCSK9: Moderate significance as a secondary marker.
- LPA: Less impactful in long-term studies.
- Environmental Interactions: PM2.5 exposure, oxidative stress, poor air quality, diet, and low physical activity exacerbate CAD progression
